# Implicit and explicit attitudes toward people with physical disabilities among clinicians, rehabilitation assistants, and other occupations: A comparative study

**DOI:** 10.1101/2025.02.22.25322346

**Authors:** Matthieu P. Boisgontier

## Abstract

**Objective:** Ableism is rooted in attitudes that influence behaviors and decisions toward people with disabilities. To assess whether these attitudes vary by occupation, we compared the preferences for people with or without physical disabilities between clinicians, rehabilitation assistants, and individuals in other professions.

**Methods:** Data from 213,191 participants, collected online through Project Implicit, were analyzed, including 6445 clinicians, 3482 rehabilitation assistants, and 203,264 individuals in other occupations. Implicit attitudes were assessed using D-scores derived from the Implicit Association Test. Explicit attitudes were assessed using a Likert scale. Regression models were conducted to examine the association between occupation groups and attitudes toward people with and without physical disabilities, while controlling for demographic variables.

**Results:** Clinicians and rehabilitation assistants showed both an implicit and explicit preference for people without physical disabilities. Implicit attitudes of clinicians and rehabilitation assistants were equivalent to those in other occupations. Compared to other occupations, clinicians had less favorable explicit attitudes toward people with physical disabilities, whereas rehabilitation assistants had more favorable ones. Older age, male sex, and no personal experience of disability were associated with less favorable attitudes toward people with physical disabilities. Associations with education, race, geographic region, and year of data collection were also observed.

**Conclusions:** This study provides evidence suggesting the persistence of ableist attitudes among clinicians and rehabilitation assistants. Moreover, implicit attitudes were similar to those of other occupations, while explicit attitudes of clinicians were even slightly less favorable.

**Impact:** Our findings suggest that despite ongoing educational shifts toward more inclusive approaches, the longstanding framework of disability as an abnormality to be normalized may still affect healthcare practitioners. This underscores the need for continued efforts to address ableism not only in healthcare, but throughout society, by promoting disability-inclusive education and training.

## INTRODUCTION

An attitude is "a psychological tendency that is expressed by evaluating a particular entity with some degree of favor or disfavor".^1^ Explicit attitudes can be reported using self-report instruments such as questionnaires, and their activation can be consciously controlled.^2^ In contrast, implicit (or automatic)^3^ attitudes, typically assessed using reaction time tasks, are traces of past experience that remain introspectively unidentified.^4^ Attitudes not only predict behavior,^5,6^ they also shape interpersonal interactions and influence decision making in professional contexts.^7,8^

In healthcare, attitudes toward people with disabilities are considered a primary measurable indicator of ableism, defined as "a set of assumptions and practices promoting the differential or unequal treatment of people because of actual or presumed impairment or disability that privileges one way of being based on normative expectations of capability and independence".^9^ Understanding implicit and explicit attitudes toward people with disabilities in healthcare practitioners is essential to identifying potential biases in care. This understanding is particularly important given the historical and prevailing view of disability through a deficit framework.^10^ This framework views disability as an abnormality that needs to be normalized to conform to societal ideals of "normalcy".^10^ This view has been embedded in healthcare practice for decades, particularly in rehabilitation professions, where a biomechanical approach to correcting deficits is foundational.^11^ Although this deficit framework has been criticized for overlooking inclusion and accessibility,^10,11^ whether healthcare practitioners’ attitudes still reflect this framework remains unclear.

This deficit-based framework aligns with what is commonly referred to as the medical or biomedical model of disability, which contrasts with the social model that defines disability as the result of structural, institutional, and attitudinal barriers rather than inherent deficits.^12,13^ While the social model has been instrumental in shifting approaches to disability toward structural barriers, it has also been criticized for undervaluing the role of physical and psychological realities.^14^ More integrated models propose that disability arises from the interaction between bodily differences and social context.^10,14,15^ This perspective is exemplified by the World Health Organization’s (WHO) International Classification of Functioning, Disability and Health (ICF).^16^ The ICF offers a biopsychosocial framework that considers disability as an umbrella term for the consequences of health conditions, including impairments of body structures or functions, activity limitations, and restrictions in participation.^16^ In these integrated models, attitudes toward disability may function as a key mechanism through which cultural norms and institutional structures influence access to care, clinical judgments, and health-related outcomes.

A systematic review investigating explicit attitudes toward people with disabilities in healthcare students and practitioners reported mixed results.^17^ Some studies found positive attitudes in occupational therapy students and professionals.^18,19^ Other studies reported more favorable attitudes in physical therapists compared to schoolteachers^20^ or the general population,^21^ and in occupational therapy students compared to business students.^22^ However, other studies found less favorable explicit attitudes toward people with disabilities in healthcare students, including nursing, medical, and rehabilitation students, compared to the general population.^23^ Negative explicit attitudes toward children with disabilities were also observed in nursing students and professionals.^24^ In addition, dental surgery assistants and dental students showed less favorable explicit attitudes toward people with disabilities than psychology students,^25^ and no statistical difference was found between occupational therapy students and business students.^26^

Recently, several studies have used data collected by Project Implicit between 2006 and 2021 to examine implicit and explicit attitudes using the Implicit Association Test (IAT) for general disability (Suppl. Fig. 1).^27–29^ One of these studies focused on healthcare professionals (n = 25,006), including clinicians, occupational and physical therapy assistants, nursing and home health assistants, technologists, technicians, and other healthcare support personnel.^27^ Results showed a slight explicit preference (Likert score = 4.41 ± 0.90) and a moderate implicit preference (D-score = 0.54 ± 0.43) for people without disabilities. A follow-up analysis of 6113 occupational and physical therapy assistants found similar results (Likert score = 4.29 ± 0.80; D-score = 0.51 ± 0.44).^28^ Compared to participants from the general population (n = 8,544; Likert score = 4.06 ± 1.17; D-score = 0.45 ± 0.43),^29^ these results suggest that healthcare professionals, including rehabilitation assistants, may have less favorable implicit and explicit attitudes toward people with disabilities. However, methodological differences across studies limit our ability to draw robust conclusions from such cross-study comparisons. Although one study compared nursing and home health assistants to individuals in other occupations using the same dataset,^30^ no research to date has used this approach to compare attitudes across groups of healthcare practitioner closely related to physical disability and the general public. Moreover, previous studies have often grouped different types of disabilities together, despite emerging evidence showing that different types of disability elicit varying levels of bias.^31^ This evidence underscores the importance of assessing attitudes toward specific conditions such as physical disabilities.

The present study addresses these gaps by exclusively focusing on attitudes toward physical disabilities and by comparing these attitudes among clinicians (e.g., medical doctors, physical therapists, nurses), rehabilitation assistants, and the general population within a single, harmonized dataset. We distinguished between clinicians, who diagnose and treat patients, and rehabilitation assistants, who support clinicians in delivering therapy, to reflect the possibility that differences in education, training, and professional roles may influence attitudes toward physical disability. Finally, studies have identified factors that shape their attitudes toward people with disabilities. In healthcare professionals, these factors include younger age^27^, female sex^23,27,32–35^, white race^27,33^, personal experience with disability (e.g., having friends, acquaintances, or family members with disabilities or having a disability oneself),^19,22,27,33,34,36–38^ and country of residence.^39^ Therefore, we explored whether sex, age, personal experience of disability, education level, geographic region, race, and year of data collection were associated with implicit and explicit attitudes toward people with physical disabilities.

## METHODS

### Participants

This study is based on the physical disability IAT dataset collected from 2022 to 2024 on the Project Implicit demonstration website (https://implicit.harvard.edu/implicit/selectatest.html) and made available on the Open Science Framework (OSF) under the CC0 1.0 Universal License.^40^ The website allows any adult aged 18 years or older to participate and measure their implicit and explicit attitudes toward people with and without physical disabilities. Participants also answered demographic questions (e.g., age, sex, race, country of residence). They were informed that their data, without directly identifying information, would be made publicly available for research purposes. Project Implicit was approved by the Institutional Review Board for the Social and Behavioral Sciences at the University of Virginia, USA, and the current study was approved by the University of Ottawa Research Ethics Board (H-02-25-11349), Canada.

### Implicit Association Test for Physical Disability

#### Assessment of Implicit Attitudes

The physical disability IAT is designed to assess implicit attitudes toward people with and without physical disabilities. In other words, this test measures the strength of automatic associations between the target concepts (i.e., people with vs. without physical disabilities) and evaluative attributes (i.e., good vs. bad). The underlying principle is that participants respond more quickly when strongly associated categories share the same response key, reflecting implicit associations.

#### Procedures

Participants completed a series of categorization tasks (Figure 1), totaling 180 trials, in which they sorted words and images appearing on a computer screen into groups by pressing designated keys on a keyboard. The categories appeared on the left and right sides of the screen, and participants were instructed to press the "E" key if the presented stimulus belonged to the left-side category and the "I" key if it belonged to the right-side category. Participants were asked to respond as quickly and accurately as possible. If a participant placed a stimulus in the incorrect category, a red "X" appeared on the screen, and the correct response had to be selected before proceeding to the next trial. Participants performed seven sequential blocks (Fig. 1A): (1) Participants categorized 12 images (Fig. 1B) of people with or without physically disabilities into the respective categories: "physically disabled people" and "physically abled people". (2) Participants categorized 16 words (Fig. 2C) into evaluative attribute categories (good vs. bad). (3) The disability and attribute categories were paired for 20 trials. For example, "physically disabled people" and "good" shared the same response key, while "physically abled people" and "bad" shared the other key. (4) The third block was repeated with 40 additional trials. (5) Similar to the first block of 20 trials but "physically disabled people" and "physically abled people" switched sides. (6) Similar to the third block of 20 trials but with a different pairing (e.g., "physically disabled people" and "bad" shared the same response key, while "physically abled people" and "good" shared the other key). (7) The sixth block was repeated with 40 additional trials. Before each block, participants were provided detailed on-screen instructions, explaining the category pairing for the upcoming block and emphasizing the need for speed and accuracy. The same 6 images were used for each target concept across series (Fig. 1B). For each series, a set of 8 words was randomly selected from a set of 16 words for each attribute (Fig. 1C).

**Figure 1.**
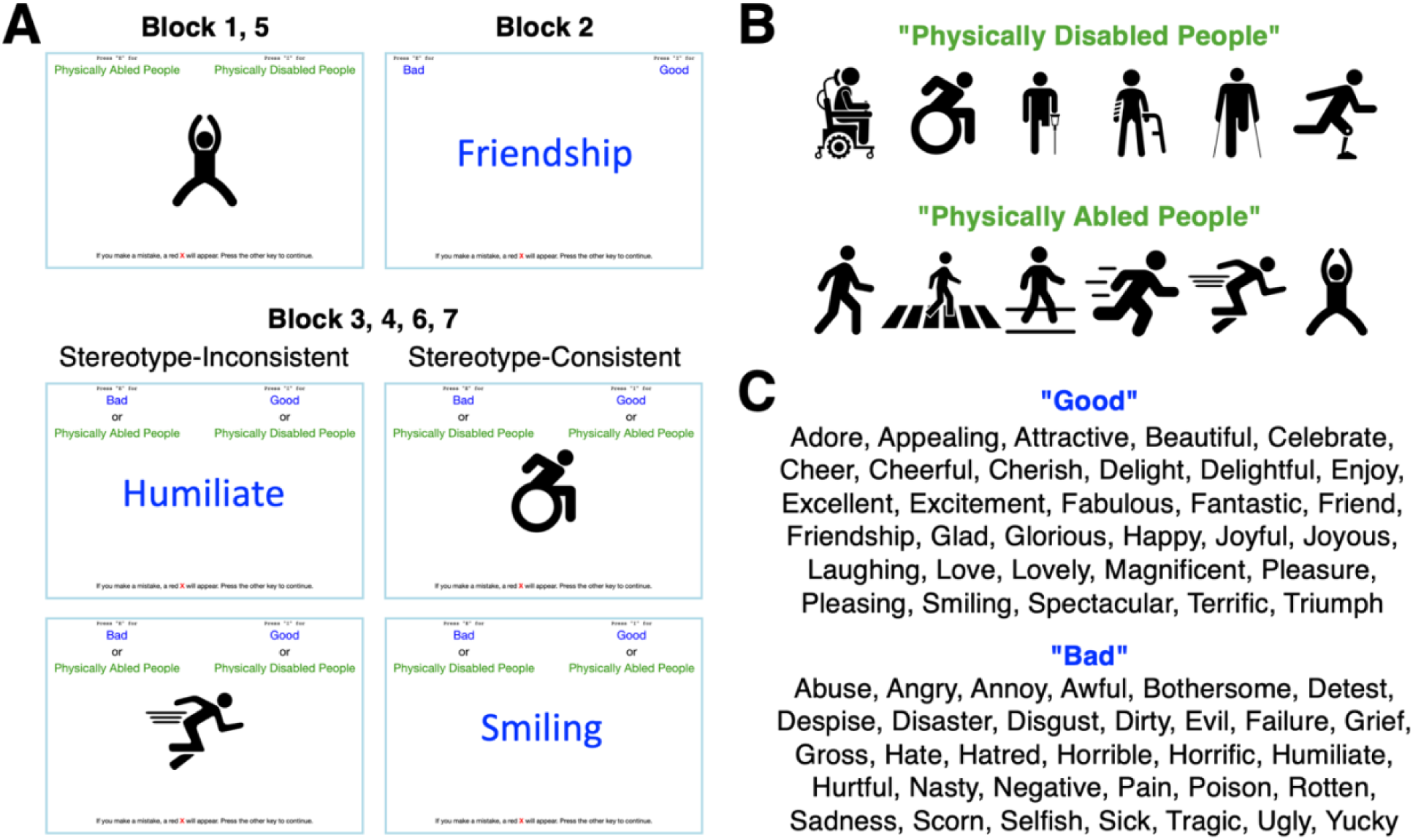
Implicit Association Test for physical disability. (A) Example screenshots from the seven trial blocks. Block 1 and 5 involved categorizing images of physically abled and physically disabled people. Block 2 involved categorizing evaluative words as "good" or "bad". Blocks 3, 4, 6, and 7 combined the target categories and evaluative attributes to form stereotype-consistent or stereotype-inconsistent pairings. (B) Images used for the target concepts ("physically disabled people" and "physically abled people"). (C) Words used for the evaluative attributes ("good" and "bad"). For each evaluative attribute, a set of 8 words was randomly selected before the start of each series.

**Figure 2.**
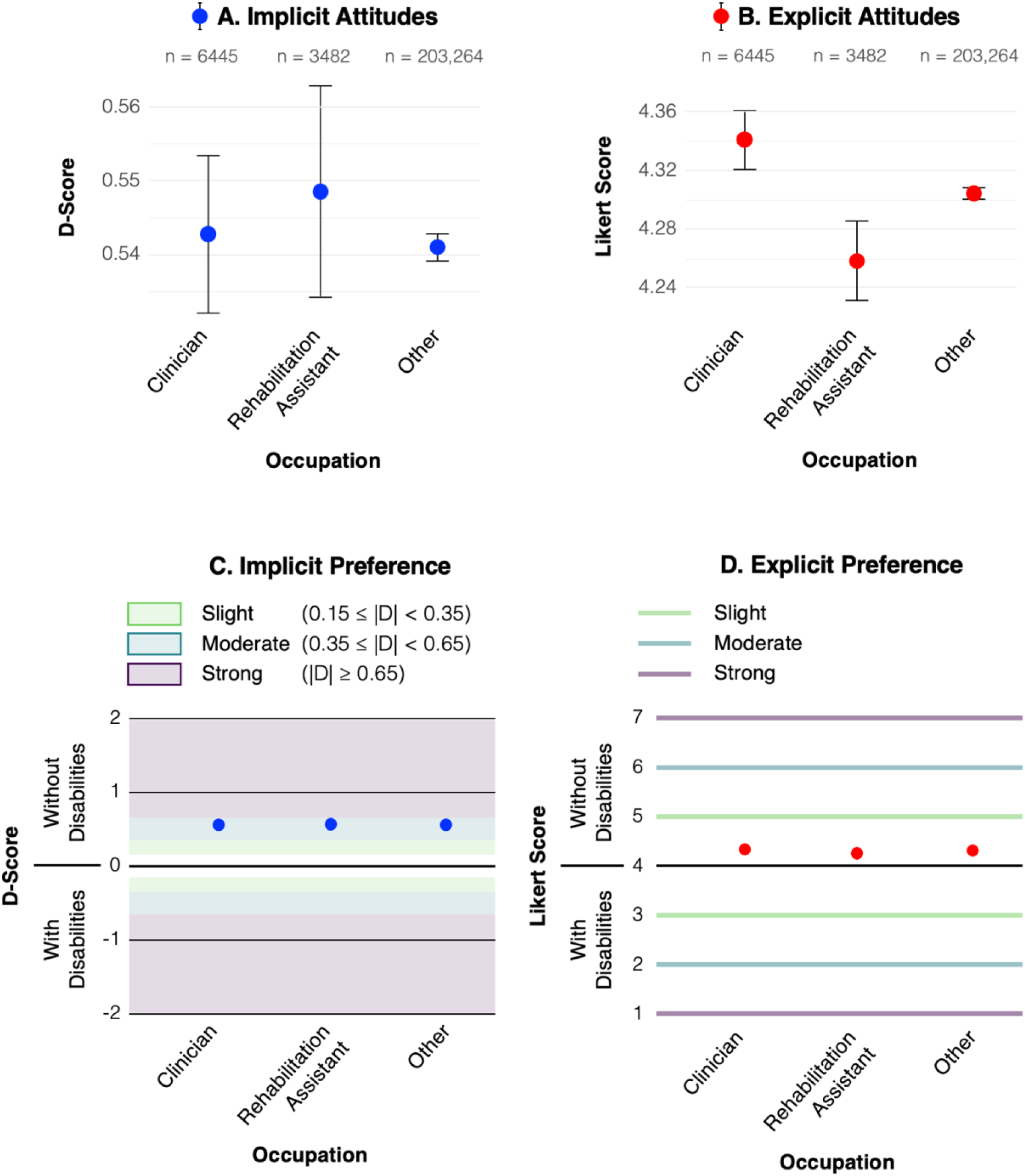
Estimated implicit (A) and explicit (B) attitudes toward physical disability across occupation groups. D-scores between 0.35 and 0.65 indicate a moderate implicit preference (C). On the 7-point Likert scale used to test explicit attitudes (D), a score of 4 was associated with "I like physically disabled people and physically abled people equally". And a score of 5 was associated with "I slightly prefer physically abled people to physically disabled people". Points represent model-based estimated means from the linear regressions adjusting for age, sex, explicit attitudes (A), implicit attitudes (B), personal experience of disability, education level, geographic region, and race. Error bars indicate 95% confidence intervals.

### Outcome Variables

#### Implicit Attitudes

Implicit attitudes toward people with and without physical disabilities were assessed using the D-score measure,^41^ which is based on participants’ performance on blocks 3, 4, 6, and 7 of the physical disability IAT. This measure divides the difference between the mean response latency on the stereotype-consistent trials (i.e., "physically disabled people" paired with "bad" and "physically abled people" paired with "good") and the mean response latency on the stereotype-inconsistent trials (i.e., "physically disabled people" paired with "good" and "physically abled people" paired with "bad") by the standard deviation of all the latencies across the four blocks:

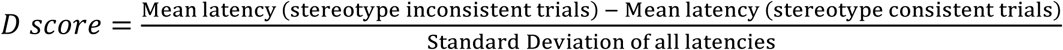

Error trials were included. Missing and incomplete data were handled using standard procedures established by Project Implicit. Only participants who completed the full IAT task and provided responses to the demographic questions were included in the public dataset. Trials with response latencies below 400 ms and above 10,000 ms were excluded to reduce the influence of random or disengaged responses, and participants with more than 10% of trials below 300 ms were excluded to ensure data validity.^41^ Moreover, participants with missing data on any of the variables used in the regression models were excluded via listwise deletion.

D-scores typically range from about –2 to 2. A positive D-score indicates that participants responded faster on stereotype-consistent trials than on stereotype-inconsistent trials, reflecting an implicit preference for people without physical disabilities. A negative D-score indicates the opposite, reflecting an implicit preference for people with physical disabilities. D-scores were interpreted as follows: no implicit preference (|D| < 0.15), slight implicit preference (0.15 ≤ |D| < 0.35), moderate implicit preference (0.35 ≤ |D| < 0.65), and strong implicit preference (|D| ≥ 0.65) (Fig. 2C).

### Explicit Attitudes

Explicit attitudes were assessed using a 7-point Likert scale in which participants rated their preference for people with or without physical disabilities. A score of 1 indicated a strong preference for people with disabilities, 4 indicated no preference, and 7 indicated a strong preference for people without disabilities. Specifically, this measure was based on the question "Which statement best describes you?", with response options as follows: (1) "I strongly prefer physically disabled people to physically abled people", (2) "I moderately prefer physically disabled people to physically abled people", (3) "I slightly prefer physically disabled people to physically abled people", (4) "I like physically disabled people and physically abled people equally", (5) "I slightly prefer physically abled people to physically disabled people", (6) "I moderately prefer physically abled people to physically disabled people", and (7) "I strongly prefer physically abled people to physically disabled people" (Fig. 2D).

### Explanatory Variables

#### Occupation

Participants’ occupation was determined based on their response to the item: "Please select the most appropriate occupation category". Participants who selected "Healthcare – Diagnosing and treating practitioners" (e.g., medical doctors, physical therapists, nurses, dentists) were categorized as clinicians. Participants who selected "Healthcare – Occupational and physical therapist assistants" were categorized as rehabilitation assistants. These two categories were selected because they represent healthcare professions directly involved in diagnosis, treatment planning, or rehabilitation, areas particularly relevant to the care of people with physical disabilities. The other healthcare-related categories (e.g., technologists, technicians, nursing assistants, and home health assistants) were included in the "other occupations" group due to less direct involvement in physical disability care. All the participants who selected non-healthcare categories were also classified under "other occupations". A complete list of occupation categories is available in Suppl. List 1.

#### Age

Age was treated as a continuous variable, which was calculated by subtracting the year of birth from the year of data collection. Since the study focused on occupational differences, participants younger than 20 and older than 70 were excluded to restrict the sample to the working-age population.

#### Sex

Participants’ sex was determined by their answer to the question "What sex were you assigned at birth, on your original birth certificate?"

#### Personal Experience of Disability

Personal experience of disability was derived from two questions: "Do you have a disability or learning difficulty?" and "Do you have a close friend or family member with a disability or learning difficulty?" Responses to these questions were combined into a new categorical variable. People who answered "yes" to either question were classified as having personal experience of disability, while those who answered "no" to both questions were classified as having no personal experience of disability.

### Education Level

Education level was categorized into three groups based on participants’ answer to the question: "Please indicate the highest level of education that you have completed". Participants who selected "elementary school", "junior high or middle school", "some high school”, or "high school graduate" were categorized as having primary or secondary education. Participants who selected "some college", "associate’s degree", or "bachelor’s degree" were categorized as having college or undergraduate education. Participants who selected "some graduate school", "master’s degree", "M.B.A.", "J.D.", "M.D.", "Ph.D.", or "other advanced degree" were categorized as having graduate or postgraduate education.

### Geographic Region

Geographic region of residence was derived from the question: "What is your country/region of primary residence?" The numeric country codes from participants’ responses were merged with the Standard Country or Area Codes for Statistical Use.^42^ The resulting categorical variable included six geographic regions: Africa, Asia, Europe, Latin America and the Caribbean, Northern America, and Oceania.

### Race

Race was determined by the question: "What is your race or ethnicity?" Participants selected from eight predefined categories: "American Indian or Alaska Native", " Asian", "Black or African American", "Hispanic", "Middle Eastern", "Pacific Islander", "Multiracial, other, or unknown", "White".

### Year

The year of data collection was included in the models as a continuous variable.

### Statistical Analyses

#### Main Analyses

All analyses were conducted in R version 4.4.1.^43^ Linear regression models were fitted using the lm() function to examine the relationship between attitudes and occupation. The outcome was either the D-score (implicit attitudes) or the Likert score (explicit attitudes). Occupation was the exposure. Age, sex, personal experience of disability, education level, geographic region, race, year of data collection, and the other type of attitudes were used as control variables. Continuous variables were standardized. The significance level (α) was set at 0.05 for all statistical tests. Since an absence of statistical significance should not be interpreted as an absence of effect,^44^ we conducted equivalence tests to further examine the nonsignificant effect of occupation on implicit attitudes.

#### Equivalence Testing

Equivalence testing is a statistical method used to formally demonstrate that two groups do not differ by more than a specified margin. Here, this method was used to determine whether clinicians and rehabilitation assistants had implicit attitudes that were statistically equivalent to those of individuals in other occupations. We used the tsum_TOST() function of the TOSTER package^45^ to conduct two-sample equivalence tests based on Welch’s two-sample t-test approach. The equivalence bounds for the D-score were set based on a smallest effect size of interest (SESOI) of ± 0.15, consistent with the interpretation thresholds described above. The SESOI was scaled by multiplying it by the residual standard error from the multiple linear regression model.^46^

Estimated D-scores, standard errors, and sample sizes were derived from the linear regression model. The residual standard error from this model was used for the reference group (other occupations). The null hypothesis for the equivalence test was that the effect was greater than the equivalence bounds, while the alternative hypothesis was that the effect was within the equivalence bounds. Thus, a significant result would indicate that the implicit attitudes of clinicians or rehabilitation assistants are statistically equivalent to those of individuals in other occupations.

## RESULTS

### Descriptive Results

A total of 213,191 participants from three occupation groups were included in the study (Table 1): clinicians (n = 6445), rehabilitation assistants (n = 3482), and participants in other occupations (n = 203,264). Implicit attitudes were similar across occupation groups, with clinicians scoring 0.54 ± 0.44, rehabilitation assistants 0.50 ± 0.44, and participants in other occupations 0.54 ± 0.44. In terms of explicit attitudes, clinicians scored 4.38 ± 0.77, rehabilitation assistants 4.25 ± 0.73, and participants in other occupations 4.30 ± 0.82. The mean age of clinicians was 35.0 ± 12.3 years, which was older than rehabilitation assistants (28.7 ± 9.7 years) and similar to participants in other occupations (34.2 ± 12.4 years). Female participants represented 72.2% of clinicians, 83.7% of rehabilitation assistants, and 76.8% of participants in other occupations. A larger proportion of participants in all occupation groups had personal experience of disability, either themselves, a family member, or a friend with 59.0% of clinicians, 64.2% of rehabilitation assistants, and 67.9% of participants in other occupations reporting such experience. Education level varied between occupation groups. Most clinicians (79.3%) had a graduate or postgraduate degree, while 19.7% had a college or undergraduate education, and 1.0% had a primary or secondary education. Among rehabilitation assistants, 49.5% had a graduate or postgraduate degree, 46.7% had a college or undergraduate education, and 3.8% had a primary or secondary education. In other occupations, these proportions were 57.7%, 34.5%, and 7.8%, respectively. Most clinicians (89.4%) and rehabilitation assistants (90.7%) were from Northern America, with lesser representation from other regions. Regarding race, 69.6% of clinicians, 76.0% of rehabilitation assistants, and 67.3% of participants in other occupations identified as White people. Asian people were the second most represented race (8.3 to 13.9%).

**Table 1.** Descriptive characteristics of the study sample by occupation group.

| **Exposures** | **Clinicians**<br>(n = 6445) | **Rehabilitation Assistants**<br>(n = 3482) | **Other Occupations**<br>(n = 203,264) |
| --- | --- | --- | --- |
| | Mean ± SD or<br>Count (%) | Mean ± SD or<br>Count (%) | Mean ± SD or<br>Count (%) |
| **Implicit Attitudes** | $0.54 \pm 0.44$ | $0.50 \pm 0.44$ | $0.54 \pm 0.44$ |
| **Explicit Attitudes** | $4.38 \pm 0.77$ | $4.25 \pm 0.73$ | $4.30 \pm 0.82$ |
| **Age** | $35.0 \pm 12.3$ | $28.7 \pm 9.7$ | $34.2 \pm 12.4$ |
| **Female Participant** | 4650 (72.2) | 2914 (83.7) | 156,161 (76.8) |
| **Personal Experience of Disability** | 3803 (59.0) | 2236 (64.2) | 138,019 (67.9) |
| **Education Level** |  |  |  |
| Primary or Secondary | 62 (1.0) | 133 (3.8) | 15,920 (7.8) |
| College or Undergraduate | 1270 (19.7) | 1625 (46.7) | 70,085 (34.5) |
| Graduate or Postgraduate | 5113 (79.3) | 1724 (49.5) | 117,259 (57.7) |
| **Geographic Region** |  |  |  |
| Africa | 47 (0.7) | 17 (0.5) | 2694 (1.3) |
| Asia | 221 (3.4) | 90 (2.6) | 10,422 (5.1) |
| Europe | 183 (2.8) | 90 (2.6) | 9501 (4.7) |
| Latin America and the Caribbean | 126 (2.0) | 51 (1.5) | 6614 (3.3) |
| Northern America | 5764 (89.4) | 3159 (90.7) | 170,761 (84.0) |
| Oceania | 104 (1.6) | 75 (2.1) | 3272 (1.6) |
| **Race** |  |  |  |
| American Indian or Native People | 37 (0.6) | 19 (0.6) | 2244 (1.1) |
| Asian People | 895 (13.9) | 289 (8.3) | 17,026 (8.4) |
| Black or African American People | 332 (5.1) | 172 (4.9) | 18,478 (9.1) |
| Hispanic People | 238 (3.7) | 135 (3.9) | 12,824 (6.3) |
| Middle Eastern People | 84 (1.3) | 23 (0.7) | 1323 (0.6) |
| Multiracial, Other, or Unknown | 358 (5.5) | 184 (5.3) | 13,851 (6.8) |
| Pacific Islander People | 17 (0.3) | 12 (0.3) | 759 (0.4) |
| White People | 4484 (69.6) | 2648 (76.0) | 136,759 (67.3) |

### Statistical Results

#### Implicit Attitudes – All Occupations

Results from the linear regression model showed no evidence suggesting that clinicians (b = 0.0017 [95% CI: -0.0090 to 0.0125]; *P* = .752) or rehabilitation assistants (b = 0.0075 [95% CI: -0.0069 to 0.0218]; *P* = .307) differed from individuals in other occupations on implicit attitudes (Fig. 2A; Fig. 3A). This absence of significant differences, combined with a significantly positive intercept (b = 0.5164 [95% CI: 0.5068 to 0.5261]; *P* < 2.0 × 10^-16^) representing the mean implicit attitude score for individuals in other occupations, indicated that all occupational groups had a D-score significantly greater than zero, i.e., an implicit preference for people without physical disabilities (Fig. 2C).

**Figure 3.**
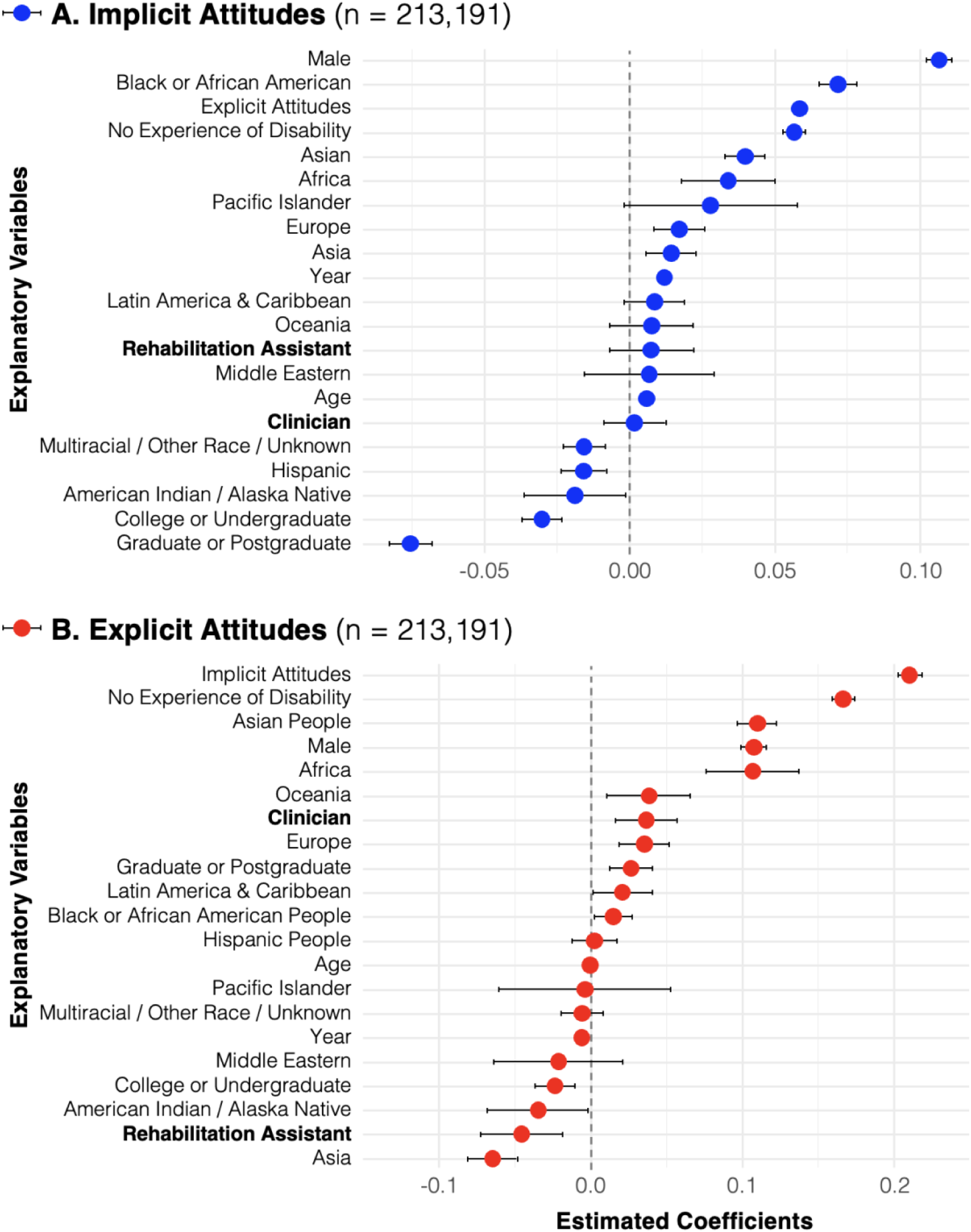
Regression coefficients from the linear models examining the association between explanatory variables with less favorable (negative coefficient) or more favorable (positive coefficient) implicit (A) and explicit (B) attitudes toward people with physical disabilities in all participants, relative to the reference categories. The reference categories are "other occupation", "female", "personal experience of disability", "Northern America", "White race", and "primary or secondary education". The figure shows the estimated coefficients (points) and 95% confidence intervals (error bars). For clarity, the continuous variables (i.e., age, explicit attitudes, implicit attitudes, year of data collection) are presented in their original units.

Age was associated with implicit attitudes toward people with physical disabilities with a coefficient regression of b = 0.0709 ([95% CI:0.0690 to 0.0728]; *P* < 2.0 × 10^-16^), corresponding to a 0.3 increase in D-score over 50 years. Male participants showed less favorable implicit attitudes than female participants (b = 0.1066 [95% CI: 0.1022 to 0.1110]; *P* < 2.0 × 10^-16^). Personal experience of disability was associated with more favorable implicit attitudes (b = 0.0568 [95% CI: 0.0528 to 0.0607]; *P* < 2.0 × 10^-16^). Higher education was associated with more favorable implicit attitudes. Specifically, participants with a college or undergraduate education (b = -0.0303 [95% CI: -0.0374 to -0.0232]; *P* < 2.0 × 10^-16^) and those with graduate or postgraduate education (b = -0.0756 [95% CI: -0.0831 to -0.0682]; *P* < 2.0 × 10^-16^) had more favorable attitudes than participants with primary or secondary education. Regionally, participants from Africa (b = 0.0339 [95% CI: .0177 to 0.0500]; *P* = 3.9 × 10^-5^), Europe (b = 0.0170 [95% CI: 0.0082 to 0.0258]; *P* = 1.5 × 10^-4^), and Asia (b = 0.0142 [95% CI: 0.0055 to 0.0229]; *P* = 1.3 × 10^-3^) had less favorable implicit attitudes than participants from other regions. By race, Black or African American (b = 0.0719 [95% CI: 0.0654 to 0.0785]; *P* = < 2.0 × 10^-16^) and Asian (b = 0.0399 [95% CI: 0.0330 to 0.0468]; *P* < 2.0 × 10^-16^) participants showed less favorable implicit attitudes, whereas American Indian or Alaska Native (b = -0.0190 [95% CI: -0.0366to -0.0013]; *P* = .035), Hispanic (b = - 0.0158 [95% CI: -0.0236 to -0.0080]; *P* = 7.4 × 10^-5^), and multiracial people or from another race (b = -0.0157 [95% CI: -0.0231 to -0.0084]; *P* = 2.8 × 10^-5^) showed more favorable implicit attitudes. Implicit attitudes toward people with physical disabilities became more unfavorable over the years of data collection (b = 0.0099 [95% CI: 0.0080 to 0.0118]; *P* < 2.0 × 10^-16^). Explicit attitudes were positively associated with implicit attitudes (b = 0.0483 [95% CI: 0.0464 to 0.0501]; *P* < 2.0 × 10^-16^). The other effects were not significant (Table 2)

**Table 2.** Association between occupation and implicit attitudes toward people with physical disabilities. Results of the linear regression model examining the relationship between occupation and implicit attitudes toward physical disability, as measured by the Implicit Association Test (IAT). The model adjusts for sex, age, explicit attitudes, personal experience of disability, education, geographic region, race, and year of data collection. Estimates are presented as regression coefficients (b-values) with 95% confidence intervals (CIs). The reference groups for categorical variables are indicated in parentheses. Asterisks indicate statistical significance (\**P* < .05, \*\**P* < .01, \*\*\**P* < .001).

| IMPLICIT ATTITUDES – All occupations (N = 213,191) |  |  |
| --- | --- | --- |
| Exposure | b-value (95% CI) | P-value |
| (Intercept) | 0.532 (0.525, 0.539) | < 2e<sup>-16</sup>*** |
| Occupation (ref.: Other occupation) |  |  |
| Clinician | 0.002 (-0.009, 0.012) | .752 |
| Rehabilitation Assistant | 0.007 (-0.007, 0.022) | .307 |
| Sex (ref.: Female) | 0.107 (0.102, 0.111) | < 2e<sup>-16</sup>*** |
| Age (continuous) | 0.071 (0.069, 0.073) | < 2e<sup>-16</sup>*** |
| Explicit Attitudes (continuous) | 0.048 (0.046, 0.050) | < 2e<sup>-16</sup>*** |
| No personal experience of disability (ref.: Experience) | 0.057 (0.053, 0.061) | < 2e<sup>-16</sup>*** |
| Education (ref.: primary / secondary) |  |  |
| College / Undergraduate | -0.030 (-0.037, -0.023) | < 2e<sup>-16</sup>*** |
| Graduate / Postgraduate | -0.076 (-0.083, -0.068) | < 2e<sup>-16</sup>*** |
| Geographic Region (ref.: Northern America) |  |  |
| Africa | 0.034 (0.018, 0.050) | 3.9e<sup>-5</sup>*** |
| Asia | 0.014 (0.006, 0.023) | 1.4e<sup>-3</sup>** |
| Europe | 0.017 (0.008, 0.026) | 1.5e<sup>-4</sup>*** |
| Oceania | 0.008 (-0.007, 0.022) | .307 |
| Latin America & the Caribbean | 0.009 (-0.002, 0.019) | .106 |
| Race (ref.: White People) |  |  |
| Asian People | 0.040 (0.033, 0.047) | < 2e<sup>-16</sup>*** |
| Black or African American People | 0.072 (0.065, 0.078) | < 2e<sup>-16</sup>*** |
| Hispanic People | -0.016 (-0.024, -0.008) | 7.4e<sup>-5</sup>*** |
| Middle Eastern People | 0.007 (-0.016, 0.029) | .556 |
| American Indian or Alaska Native People | -0.019 (-0.037, -0.001) | 3.5e<sup>-2</sup>* |
| Multiracial, Other, or Unknown Race People | -0.016 (-0.023, -0.008) | 2.8e<sup>-5</sup>*** |
| Pacific Islander People | 0.028 (-0.002, 0.058) | 6.7e<sup>-2</sup> |
| Year of Data Collection | 0.010 (0.008, 0.012) | < 2e<sup>-16</sup>*** |

Residual standard error: 0.4271 on 213169 degrees of freedom. Multiple R-squared: 0.06158, adjusted R-squared: 0.06148. F-statistic: 666.1 on 21 and 213169 degrees of freedom, p-value: < 2.0e-16

The linear regression model explained 6.1% of the variance in implicit attitudes (adjusted R² = 0.061) with a significant fit (F(21,213169) = 666.71; *P* < 2.0 × 10^-16^), and a residual standard error of 0.4271.

### Equivalence Testing

To assess the equivalence of implicit attitudes between occupation groups, we conducted two equivalence tests with a SESOI of ± 0.15, scaled by the residual standard error from the linear regression model (0.4271), yielding equivalence bounds of ± 0.0642 in Cohen’s d. These bounds correspond to ± 0.0194 on the raw D-score scale.

The equivalence test comparing implicit attitudes between clinicians and individuals in other occupations was significant (t(205211.16) = -18.60, *P* < 2.0 × 10^-16^). The 90% CI for the difference in implicit attitudes between clinicians and people in other occupations ranged from 0.0002 to 0.0033. Since this difference fell within the equivalence bounds of ± 0.0194, the implicit attitudes of clinicians and those in other occupations were considered equivalent.

The equivalence test comparing implicit attitudes between rehabilitation assistants and individuals in other occupations was significant (t(206743.97) = -12.47; *P* < 2.0 × 10^-16^). The 90% CI for the difference in implicit attitudes between rehabilitation assistants and people in other occupations ranged from 0.0059 to 0.0091. Since this difference fell within the equivalence bounds, the implicit attitudes of rehabilitation assistants and those in other occupations were considered equivalent.

### Explicit Attitudes – All Occupations

Clinicians showed less favorable explicit attitudes toward people with physical disabilities than other occupations (b = 0.0364 [95% CI: 0.0161 to 0.0568]; *P* = 4.6 × 10^-4^), while rehabilitation assistants showed more favorable explicit attitudes (b = -0.0459 [95% CI: -0.0731 to -0.0187]; *P* = 9.4 × 10^-4^) (Fig. 2B, Fig. 3B). To test whether explicit attitudes were significantly greater than a Likert score of 4 representing "I like physically disabled people and physically abled people equally", we re-leveled the model to set the group with the lowest explicit attitude (rehabilitation assistants) as the reference group. Results of this model showed that the intercept, which represents the mean explicit attitude score for rehabilitation assistants, was estimated at 4.172 with a standard error of 0.0154. To determine whether this value was significantly above 4, a one-sample t-test was conducted by dividing the difference between the intercept and 4 by its standard error, yielding a t-value of 11.16 (*P* < 2.0 × 10^-16^). This result confirmed that explicit attitudes in rehabilitation assistants were significantly greater than 4. Since clinicians (b = 0.0823, [95% CI: 0.0488 to 0.1158]; *P* = 1.4 × 10^-6^) and participants in other occupations had significantly higher explicit attitude scores than rehabilitation assistants, it follows that all occupational groups had an explicit preference for people without physical disabilities (Fig. 2D).

Male participants showed less favorable explicit attitudes toward people with physical disabilities than female participants (b = 0.1076 [95% CI: 0.0992 to 0.1159]; *P* < 2.0 × 10^-16^). Participants with personal experience of disability reported more favorable explicit attitudes (b = 0.1665 [95% CI: 0.1590 to 0.1740]; *P* < 2.0 × 10^-16^). Education effects were mixed: college or undergraduate education was associate with more favorable explicit attitudes (b = -0.0238 [95% CI: -0.0372 to -0.01041]; *P* = 4.9 × 10^-4^), while graduate or postgraduate education was associated with less favorable explicit attitude (b = 0.0263 [95% CI: 0.0122 to 0.0405]; *P* = 2.6 × 10^-4^). Participants in Africa (b = 0.1067 [95% CI: 0.0761 to 0.1373]; *P* = 8.1 × 10^-12^), Oceania (b = 0.0383 [95% CI: 0.0185 to 0.0519]; *P* = 6.1 × 10^-3^), Europe (b = 0.0352 [95% CI: 0.0185 to 0.0519]; *P* = 3.6 × 10^-5^), and Latin America and the Caribbean (b = 0.0210 [95% CI: 0.0013 to 0.0407]; *P* = .037) showed less favorable explicit attitudes than participants in Northern America, while participants in Asia showed more favorable explicit attitudes (b = -0.0651 [95% CI: -0.0816 to -0.0486]; *P* = 9.7 × 10^-15^). Explicit attitudes differed by race, with Asian (b = 0.1096 [95% CI: 0.0966 to 0.1227]; *P* < 2.0 × 10^-16^) and Black or African American (b = 0.0149 [95% CI: 0.0026 to 0.0273]; *P* = .018) participants showing less favorable explicit attitudes than White participants and the other racial categories, while American Indian or Alaska Native participants showed more favorable explicit attitudes (b = -0.0350 [95% CI: -0.0683 to -0.0016]; *P* = .040). Explicit attitudes toward people with physical disabilities became less unfavorable over the years of data collection (b = -0.0047 [95% CI: -0.0083 to -0.0011]; *P* = .010). Explicit and implicit attitudes were positively associated (b = 0.0927 [95% CI: 0.0892 to 0.0963]; P < 2.0 × 10^−16^). The other effects were not significant (Table 3).

**Table 3.** Association between occupation and explicit attitudes toward people with physical disabilities. Results of the linear regression model examining the relationship between occupation and explicit attitudes toward physical disability, as measured by the 7-point Likert-type scale. The model adjusts for sex, age, implicit attitudes, personal experience of disability, education, geographic region, and race. Estimates are presented as regression coefficients (b-values) with 95% confidence intervals (CIs). Asterisks indicate statistical significance (\**P* < .05, \*\**P* < .01, \*\*\**P* < .001).

| **EXPLICIT ATTITUDES – All occupations (N = 213,191)** |  |  |
| --- | --- | --- |
| **Exposure** | **b-value (95% CI)** | **P-value** |
| (Intercept) | 4.218 (4.205, 4.231) | $< 2e^{-16}$*** |
| Occupation (ref.: Other occupation) |  |  |
| Clinician | 0.036 (0.016, 0.057) | $4.59e^{-4}$*** |
| Rehabilitation Assistant | -0.046 (-0.073, -0.019) | $9.44e^{-4}$*** |
| Sex (ref.: Female) | 0.108 (0.099, 0.116) | $< 2e^{-16}$*** |
| Age (continuous) | -0.003 (-0.006, 0.001) | .169 |
| Implicit Attitudes (continuous) | 0.093 (0.089, 0.096) | $< 2e^{-16}$*** |
| No personal experience of disability (ref.: Experience) | 0.166 (0.159, 0.174) | $< 2e^{-16}$*** |
| Education (ref.: primary / secondary) |  |  |
| College / Undergraduate | -0.024 (-0.037, -0.010) | $4.91e^{-4}$*** |
| Graduate / Postgraduate | 0.026 (0.012, 0.040) | $2.62e^{-4}$*** |
| Geographic Region (ref.: Northern America) |  |  |
| Africa | 0.107 (0.076, 0.137) | $8.1e^{-12}$*** |
| Asia | -0.065 (-0.082, -0.049) | $9.7e^{-15}$*** |
| Europe | 0.035 (0.019, 0.052) | $3.6e^{-5}$*** |
| Oceania | 0.038 (0.011, 0.066) | $6.1e^{-3}$** |
| Latin America & the Caribbean | 0.021 (0.001, 0.041) | $3.7e^{-2}$* |
| Race (ref.: White People) |  |  |
| Asian People | 0.110 (0.097, 0.123) | $< 2e^{-16}$*** |
| Black or African American People | 0.015 (0.003, 0.027) | $1.8e^{-2}$* |
| Hispanic People | 0.002 (-0.012, 0.017) | .761 |
| Middle Eastern People | -0.022 (-0.064, 0.021) | .316 |
| American Indian or Alaska Native People | -0.035 (-0.068, -0.002) | $4.0e^{-2}$* |
| Multiracial, Other, or Unknown Race People | -0.006 (-0.020, 0.008) | .431 |
| Pacific Islander People | -0.004 (-0.061, 0.053) | .893 |
| Year of Data Collection | -0.005 (-0.008, -0.001) | $1.02e^{-2}$* |
| Residual standard error: 0.8088 on 213169 degrees of freedom. Multiple R-squared: 0.03364, adjusted R-squared: .03354. F-statistic: 353.4 on 21 and 213169 degrees of freedom, p-value: $< 2.0e^{-16}$ | | |

The linear regression model explained 3.4% of the variance in explicit attitudes (adjusted R² = 0.034), with a significant fit (F(21,213169) = 353.4; *P* < 2.0 × 10^-16^), and a residual standard error of 0.803.

### Attitudes in Clinicians

Among clinicians, implicit attitudes were less favorable in male participants (b = 0.1301 [95% CI: 0.1068 to 0.1534]; *P* < 2 × 10^-16^) as well as in older participants with a regression coefficient of b = 0.0719 ([95% CI: 0.0609 to 0.0829]; *P* < 2 × 10^-16^) (Fig. 4A), corresponding to a 0.3 increase in D-score over 50 years (Fig. 5A). Clinicians with no personal experience of disability showed less favorable implicit attitudes (b = 0.0460 [95% CI: 0.0245 to 0.0675]; *P* = 2.7 × 10^-5^). Implicit attitudes became more defavorable over the years of data collection (b = 0.0112 [95% CI: 0.0005 to 0.0219]; *P* = .040). Explicit and implicit attitudes were positively associated (b = 0.0546 [95% CI: 0.0441 to 0.0651]; *P* < 2 × 10^-16^) (Fig. 5B). The other effects were not statistically significant (Suppl. Table 1).

**Figure 4.**
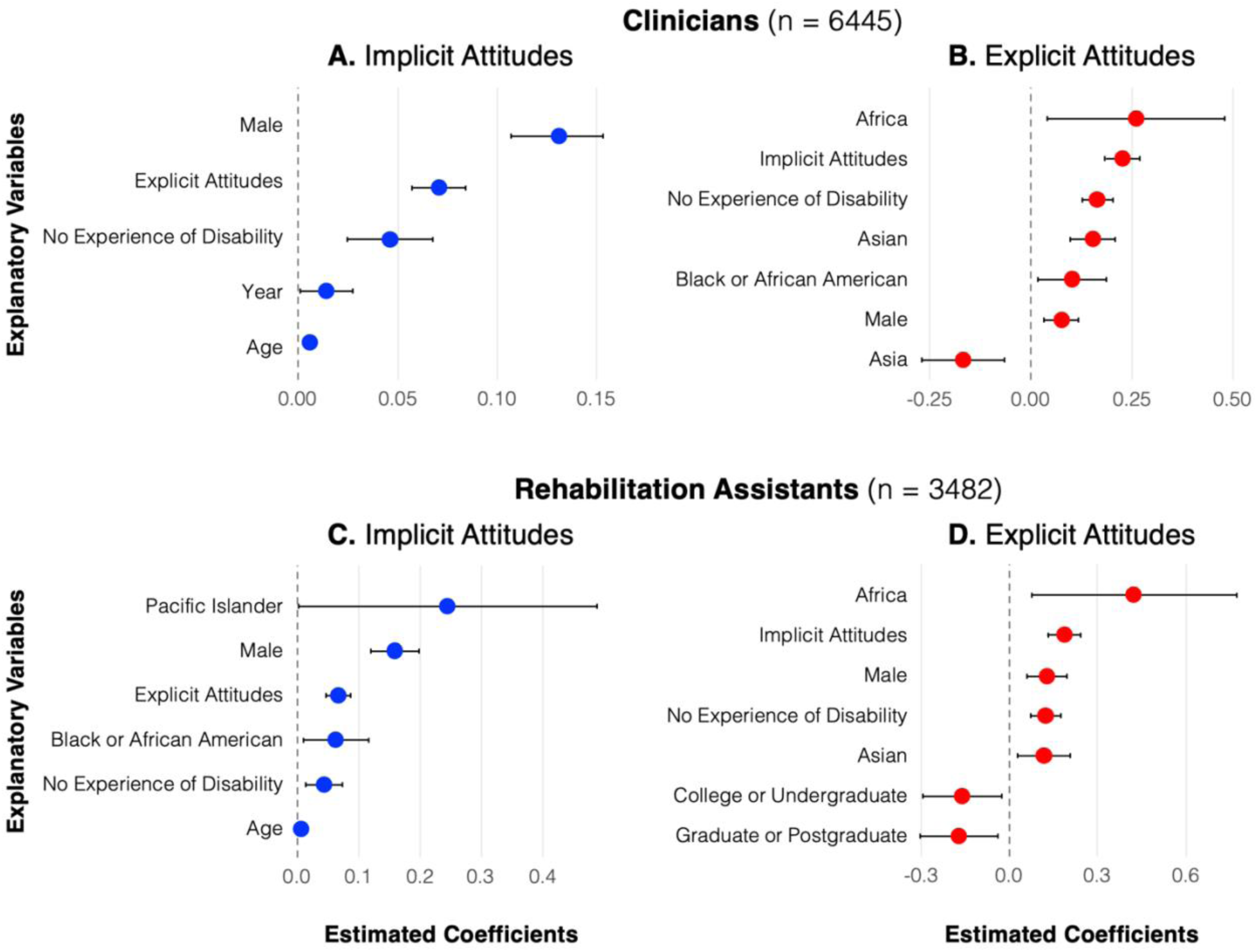
Regression coefficients from the linear models significantly associating explanatory variables with implicit (left panel) and explicit (right panel) attitudes toward people with physical disabilities, relative to the reference categories, in clinicians (top panel) and rehabilitation assistants (bottom panel). Positive coefficients indicate less favorable attitudes toward people with physical disabilities, whereas negative coefficients indicate more favorable attitudes toward people with physical disabilities. The reference categories are "female", "personal experience of disability", "Northern America", "White race", "primary or secondary education". The figure displays estimated coefficients (points) with 95% confidence intervals (error bars). For clarity, continuous variables (i.e., age, explicit attitudes, implicit attitudes, year of data collection) are presented in their original units.

**Figure 5.**
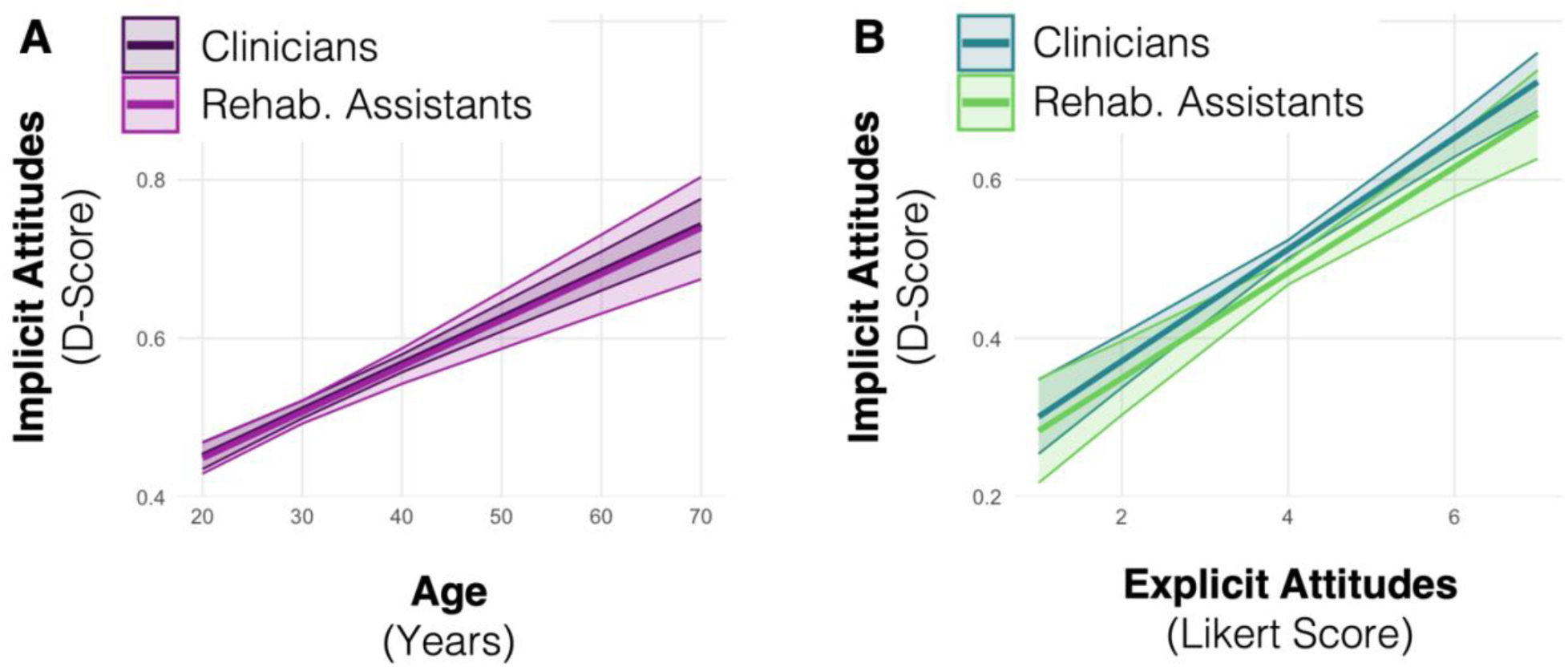
Estimated effect of age (A) and explicit attitudes (B) on the implicit preference for people without physical disabilities (positive D-score) in clinicians and rehabilitation assistants. The shaded area represents the 95% confidence interval.

Analysis of explicit attitudes showed that male clinicians had less favorable explicit attitudes toward people with physical disabilities than female clinicians (b = 0.0761 [95% CI: 0.0339 to 0.1184]; *P* = 4.1× 10^-4^) (Fig. 4B). Clinicians who reported no personal experience of physical disabilities had less favorable implicit attitudes toward people with physical disabilities than clinicians who had this experience (b = 0.1651 [95% CI: 0.1266 to 0.2035]; *P* < 2 × 10^-16^). Geographic region of residence was associated with explicit attitudes, with clinicians from Asia displaying more favorable explicit attitudes toward people with physical disabilities than people from Northern America (b = -0.1671 [95% CI: -0.2703 to -0.0638]; *P* = 1.5 × 10^-3^), and clinicians from Africa showing less favorable explicit attitudes (b = 0.2610 [95% CI: 0.0424 to 0.4797]; *P* = .019). Explicit attitudes differed by race, with Asian (b = 0.1541 [95% CI: 0.0984 to 0.2098]; *P* = 6.1 × 10^-8^) and Black or African American (b = 0.1043 [95% CI: 0.0193 to 0.1892]; *P* = .016) clinicians showing less favorable explicit attitudes toward people with physical disabilities than White clinicians. Implicit and explicit attitudes were positively associated (b = 0.0994 [95% CI: 0.0804 to 0.1185]; *P* < 2 × 10^-16^). The other effects were not statistically significant (Suppl. Table 2).

### Attitudes in Rehabilitation Assistants

Rehabilitation assistants showed less favorable implicit attitudes among male (b = 0.1590 [95% CI: 0.1196 to 0.1985]; *P* = 3.5 × 10^-15^) and older participants (b = 0.0564 [95% CI: 0.0416 to 0.0711]; *P* = 1.0 × 10^-13^) (Fig. 4C; Fig. 5A). Rehabilitation assistants with no personal experience of disability showed less favorable implicit attitudes (b = 0.0426 [95% CI: 0.0123 to 0.0729]; *P* = 5.9 × 10^-3^). Racial differences emerged, with Asian (b = 0.0619 [95% CI: 0.0089 to 0.1150]; *P* = .022) and Pacific Islander (b = 0.2449 [95% CI: 0.0013 to 0.4885]; *P* = .049) rehabilitation assistants showing less favorable explicit attitudes than White rehabilitation assistants. Implicit and explicit attitudes were positively associated (b = 0.0489 [95% CI: 0.0344 to 0.0633]; P = 3.9 × 10^-11^) (Fig. 5B). The other effects were not statistically significant (Suppl. Table 3).

In terms of explicit attitudes, male rehabilitation assistants (b = 0.1295 [95% CI: 0.0627 to 0.1963]; *P* = 1.5 × 10^-4^) and those without personal experience with disability (b = 0.1252 [95% CI: 0.0744 to 0.1761]; *P* = 1.4 × 10^-6^) had less favorable explicit attitudes (Fig. 4D). Rehabilitation assistant from Africa (b = 0.4234 [95% CI: 0.0777 to 0.7691]; *P* = .016) and Asian rehabilitation assistants (b = 0.1196 [95% CI: 0.0303 to 0.2088]; *P* = 8.6 × 10^-3^) showed less favorable explicit attitudes than those from Northern America and White rehabilitation assistants, respectively. Rehabilitation assistants with college or undergraduate education (b = -0.1679 [95% CI: -0.2986 to -0.0372]; *P* = .012) and graduate or postgraduate education (b = -0.1567 [95% CI: -0.2895 to - 0.0239]; *P* = .021) had more favorable explicit attitudes than those with primary or secondary education. Implicit and explicit attitudes were positively associated (b = 0.0833 [95% CI: 0.0587 to 0.1080]; P = 3.9× 10^-11^). The other effects were not statistically significant (Suppl. Table 4).

## DISCUSSION

### Main Findings

The present study examined implicit and explicit attitudes toward people with physical disabilities among clinicians, rehabilitation assistants, and individuals in other occupations. Results indicated that all occupational groups exhibited a moderate implicit preference for people without physical disabilities. Additionally, no significant differences in implicit attitudes were found between clinicians, rehabilitation assistants, and participants in other occupations, as confirmed by equivalence testing. Results showed a slight explicit preference for people without physical disabilities in all occupation groups, but with small differences between occupations: clinicians had less favorable explicit attitudes toward people with disabilities compared to those in other occupations, whereas rehabilitation assistants had more favorable explicit attitudes.

In clinicians and rehabilitation assistants, several demographic factors were significantly associated with implicit and explicit attitudes toward people with physical disabilities. Male participants exhibited less favorable implicit and explicit attitudes than female participants. Personal experience of disability was associated with more favorable implicit and explicit attitudes. Older participants showed less favorable implicit attitudes. Education attainment influenced explicit attitudes, with higher levels of education associated with more favorable attitudes. Residents of Africa and Asian participants showed less favorable explicit attitudes toward people with disabilities than those from the other countries.

### Comparison with the Literature

Our findings support previous research showing less favorable attitudes toward people with general disabilities in healthcare students and practitionners.^23,25^ Specifically, similar to results from prior studies in healthcare professionals,^27–29^ our findings showed a moderate implicit and slight explicit preference for people without disabilities, but with a focus on physical disabilities.

Similar to a study conducted in nursing and home health assistants,^30^ we compared implicit and explicit attitudes between healthcare professionals and individuals in other occupations but focused on clinicians and rehabilitation assistants. Consistent with the findings in nursing and home health assistants, differences in attitudes between healthcare professionals and individuals in other occupations were small or not significant, with differences raging from 0.00 to 0.05 on a D-score that typically ranges from -2 to 2, and from 0.04 to 0.09 on a 7-point Likert scale in both studies. Our small effect sizes suggest that healthcare practitioners, including clinicians and rehabilitation assistants, have only slightly different attitudes toward disability than the rest of the population. The minimal differences we found may reflect the enduring effects of decades of healthcare education that has been centered on the deficit framework.^10,11^ Our findings suggest that ongoing educational shifts toward more inclusive approaches^9^ should be pursued to reshape healthcare practitioners’ attitudes toward physical disability. Although healthcare practitioners may recognize the importance of inclusion and accessibility, the persistent influence of the deficit framework may still shape their implicit and explicit attitudes, highlighting the need for further interventions to reduce biases and, in turn, improve the quality of care provided to people with disabilities.

Our results add to the evidence that male clinicians and rehabilitation assistants have less favorable implicit and explicit attitudes toward people with disabilities than female ones.^23,27,32–35^ Our results support previous literature showing an association between age and attitudes toward people with general disabilities.^27^ However, while our results showed that older age was associated with less favorable implicit attitudes toward physical disabilities, we found no evidence suggesting an association with explicit attitudes. Supporting a large body of previous research,^19,22,27,33,34,36–38^ personal experience of disability, such as having a disability oneself, or having family members, friends, or acquaintances with a disability, was statistically significant in all the models we conducted.

### Improving the International Classification of Functioning, Disability, and Health (ICF)

The present findings should also be interpreted in light of broader theoretical models of disability. The biopsychosocial perspective embedded in the World Health Organization’s International Classification of Functioning, Disability, and Health (ICF)^16^ emphasizes that disability results from the dynamic interaction between bodily differences and the social environment. By showing moderate implicit and slight explicit preferences for nondisabled individuals across all occupational groups, including clinicians and rehabilitation assistants, our results illustrate how healthcare professionals may continue to hold less favorable attitudes toward disability. As Roush and Sharby highlight,^15^ physical therapy exemplifies the tension between the medical model focused on correcting impairments and socially oriented frameworks that prioritize autonomy, participation, and dignity. Our results suggest that, 15 years later, this tension is still ongoing in rehabilitation professions and reinforce the need for further refining an integrated, patient-centered biopsychosocial model that actively challenges ableist assumptions. Specifically, attitudes toward disability should be considered a mediator of the effect of cultural norms and institutional structures on ableist behaviors, influencing access to care, clinical decision-making, and health-related outcomes. The ICF model should more explicitly incorporate attitude changes as an essential component of equitable care in rehabilitation and other healthcare professions.

### Beyond the Implicit Association Test

The absence of association between occupation group and implicit attitudes in the present study should be considered in light of the strengths and limitations of the tool used to measure these attitudes. The IAT has robust internal consistency (α = .80)^47^ and proven useful in detecting group-level attitudes in socially sensitive contexts where explicit self-reports may be compromised by social desirability bias. For example, large-scale analyses have shown evidence of implicit race bias across millions of respondents, with marked divergences from explicit self-reports.^48^ However, the IAT’s test-retest reliability is moderate (average r = .50),^49^ suggesting that half of the variability is due to stable individual differences, while the other half is due to measurement error, situational factors, or changes in the construct being measured. Its predictive validity is small to modest, with meta-analytic correlation typically ranging from r = .09 to .28.^49,50^ This modest predictive power is partly due to extraneous influences such as recoding, familiarity, and task structure, which can affect IAT performance independently of the attitudes being measured.^8^ Although the IAT is less susceptible to deliberate faking than explicit measures, social desirability can still influence responses, sometimes at a subconscious level.^51^ To address these limitations, future studies should incorporate the IAT within a broader multimethod approach for assessing disability bias. Such an approach could include approach-avoidance tasks,^52^ explicit self-report questionnaires, behavioral observations, physiological and neural measures, as well as qualitative interviews.^53^

### Implicit Attitudes and Behavior

As mentioned above, the correlation between implicit attitudes toward disability and behavioral outcomes may be small (r = 0.09).^50^ However, evidence from studies on race-, gender- , and age-related bias suggests that even modest associations can influence clinical behavior, including diagnostic reasoning and treatment decisions.^54^ For example, physicians were found to be less likely to prescribe appropriate cardiac medications to women than to men with identical symptoms,^55^ and diagnostic certainty was significantly lower for female and younger patients, leading to reduced investigation and treatment.^56,57^ These studies reported disparities in treatment recommendations ranging from 10% to 20%, even when clinical presentations were matched.

In non-clinical domains such as hiring, implicit attitudes toward disability have been shown to predict discriminatory outcomes.^58,59^ Baert^58^ found that job applicants with disabilities received significantly 48% fewer callbacks than equally qualified applicants without disabilities. Similarly, Ameri et al.^59^ found that applicants with spinal cord injury were 26% less likely to receive a positive response from employers than matched applicants without a disability. Because both studies controlled for productivity-related concerns, the disparities are best explained by underlying bias. Taken together, these findings suggest that implicit attitudes may subtly but consistently shape spontaneous behaviors, contributing to systemic inequities in healthcare and beyond.

### Educational Strategies for Reducing Bias toward People with Disabilities

Educational efforts to improve attitudes toward people with disabilities are most effective when they combine information delivery and experiential or relational learning, particularly those involving direct or indirect contact with individuals with disabilities.^60^ These multi-component interventions consistently outperform interventions that rely on information delivery alone, because they provide emotionally salient, personalized, and socially meaningful contexts that promote empathy and challenge stereotypes.^60^

For example, healthcare students who participated in direct interaction and communication training with disabled people reported improvements in comfort, empathy, and understanding.^61^ Similarly, disability simulations, when combined with reflective debriefing, emotionally engaging documentaries, and personal narratives, enhanced adolescents’ understanding of the experiences of people with disabilities and reduced negative stereotypes and social distancing behaviors.^62^ In another study, university students enrolled in a semester-long service-learning course that culminated in a week-long therapeutic camp for children with disabilities showed significantly improved attitudes and comfort levels when interacting with disabled individuals.^63^

Morehouse and Banaji^48^ argue that meaningful and long-term improvement in implicit attitudes cannot be achieved through isolated educational sessions or one-time reflection exercises. Instead, they emphasize the importance of sustained and systemic exposure to counter-stereotypical exemplars, that is, repeated interactions with disabled people presented in positive, competent, and diverse roles. This includes not only exposure to admired individuals with disabilities but also routine representation of disabled people as professionals, leaders, caregivers, and peers. Schools, universities, and healthcare training programs have an institutional responsibility to move beyond simply teaching about explicit and implicit attitudes. These institutions must actively work to “restructure experience”, a concept that Morehouse and Banaji^48^ use to describe a sustained redesign of the learning environment to make bias-disrupting experiences habitual rather than exceptional. This involves embedding disability inclusion into curricula (e.g., through case studies, images, and literature), pedagogy (e.g., using disability-inclusive teaching practices and materials), and clinical or field placements (e.g., ensuring trainees work alongside or learn from professionals with disabilities or serve diverse patient populations). Without such environmental redesign and experience repetition, interventions are unlikely to yield lasting change in attitudes toward people with disabilities Therefore, educational systems not only need to teach about bias but also to create conditions in which inclusive interactions become the norm, ultimately transforming automatic evaluative processes and behaviors.

### Strengths and Limitations

A strength of our study is that it is the first to use the Implicit Project dataset to examine attitudes toward physical disability, whereas previous research has focused on disability in general. This focus on physical disability is important because attitudes can vary depending on the target concept, and because these attitudes are particularly relevant to rehabilitation. Another strength is the use of equivalence testing, which provides statistical evidence that implicit attitudes are equivalent across occupational groups.

Several limitations should be noted. First, due to the large-scale online data collection, we cannot rule out the possibility for selection bias (e.g., only participants interested in research or with high digital literacy may have participated). Second, the percentages of variance explained by the explanatory variables were modest. However, these variables may still be meaningful in large samples and contribute to a broader understanding of attitudes. Third, Project Implicit is a repeated cross-sectional dataset that does not track the same individuals longitudinally, which precludes analyses of within-person change over time. This limitation makes it impossible to determine causal relationships. Future research should adopt prospective longitudinal designs that follow the same healthcare trainees or professionals over time, ideally linking implicit and explicit measures to observed behaviors and environmental factors. Finally, the fact that the physical disability IAT on the Implicit Project website uses identity-first language (i.e., "physically disabled people"; "physically abled people") may be seen as a limitation because person-first language (i.e., "people with physical disabilities"; "people without physical disabilities") has traditionally been promoted as a way to reduce stigma.^64^ However, recent literature suggests that person-first language in scientific writing may actually increase rather than decrease stigma.^65^ Moreover, policies mandating the use of person-first language overlook the diverse language preferences among disabled people, including disabled researchers.^66^ Accordingly, the American Psychological Association (APA) now states that "both person-first and identity-first approaches to language are designed to respect disabled persons; both are fine choices overall".^67^

## Conclusion

This study provides evidence suggesting the persistence of ableist attitudes, particularly implicit and explicit preferences for nondisabled individuals, among clinicians and rehabilitation assistants. Moreover, implicit and explicit attitudes toward people with physical disabilities were similar between clinicians, rehabilitation assistants, and individuals in other occupations. While all occupation groups showed a moderate implicit preference for people without physical disabilities, explicit attitudes varied slightly, with clinicians showing less favorable explicit attitudes and rehabilitation assistants showing more favorable explicit attitudes than those in other occupations. Taken together, these results strongly suggest that clinicians and rehabilitation assistants are not immune to implicit and explicit biases that are unfavorable to people with physical disabilities, and that their occupation does not shield them from such attitudes. Further, these findings underscore the need for continued efforts to address ableism in healthcare by promoting disability-inclusive education and training. Specifically, professional development should target both implicit and explicit attitudes to ensure that healthcare practitioners not only recognize structural and institutional barriers but also confront their own biases toward people with physical disabilities.

## Data Availability

The R script used to analyze the data are publicly available in Zenodo (https://doi.org/10.5281/zenodo.14876870). The dataset and materials for the Implicit Association Test are available in the Project Implicit Demo Website Datasets, hosted on the Open Science Framework (https://doi.org/10.17605/OSF.IO/Y9HIQ).

https://doi.org/10.5281/zenodo.14876870

## ARTICLE INFORMATION

### Data and Code Sharing

The dataset and materials for the Implicit Association Test are available in the Project Implicit Demo Website Datasets, hosted on the Open Science Framework (OSF).^34^ In agreement with good research practices,^68^ the R scripts are available on Zenodo.^69^ This manuscript was posted before peer review on the MedRxiv preprint repository on February 26, 2025.^70^

### Disclosure

The author is an associate editor for the European Rehabilitation Journal. ChatGPT (OpenAI) and DeepL were used to refine the language and improve readability of this manuscript. No other conflicts of interest were reported.

